# The predictive value of socioeconomic status and migration background for complicated lower respiratory tract infections in primary care

**DOI:** 10.1101/2025.11.06.25339670

**Authors:** Ernst D. van Dokkum, Naomi Kraaijenbrink, Saskia Le Cessie, Martijn Sijbom, Adriënne S. van der Schoor, Leo G. Visser, Cees van Nieuwkoop, Hanneke Borgdorff

## Abstract

While socioeconomic status (SES) and migration background have been linked to complicated lower respiratory tract infections (LRTIs) in population-based studies, their predictive value in primary care remains unclear. Using routine care data from Dutch general practices (Leiden-The Hague-Zoetermeer region, n ≈ 750,000 adult patients, 2014 to 2023, excluding COVID-19 years), linked to sociodemographic and hospital claims data, we developed a multivariable logistic regression model to predict 30-day hospitalisation or death following LRTI. Among 186,094 LRTI episodes, 2.19% were classified as complicated. After adjusting for established clinical factors, SES was a strong predictor, whereas migration background was not. Patients in the lowest SES category had an adjusted odds ratio of 1.46 (95%CI: 1.31 – 1.62) for a complicated course compared to the highest. The incorporation of SES into clinical decision tools and guidelines has the potential to enhance risk-stratification of patients with LRTI in daily practice of primary care, thereby supporting more equitable care.

## Introduction

Lower respiratory tract infections (LRTIs) (infectious bronchitis, bronchiolitis, and pneumonia) represent the leading cause of infectious disease burden in Europe, including the Netherlands^1,2^. LRTIs can result in severe complications, such as respiratory failure and death, either directly or through related health consequences. Seasonal peaks in LRTI incidence during winter months place substantial strain on healthcare systems, a challenge that is expected to intensify due to an ageing population and the risk of emerging pandemics.

The Dutch healthcare system is characterised by universal coverage and broad access to primary, hospital, and specialist care, ranking among the most accessible in the European Union^3,4^. Nevertheless, significant health disparities persist in various health outcomes^5,6^, as in other European countries with highly accessible care^7-9^. Within the Dutch healthcare system, general practitioners (GPs) act as gatekeepers to hospital and specialist care^10,11^. This makes primary care the central setting for managing LRTIs, which provides a unique opportunity to study LRTIs course of disease, using routinely collected, real-world primary care data.

Coughing, often indicative of an LRTI, is among the most common reasons for GP consultations in the Netherlands^12^. Severely ill patients are typically referred to hospital care the same day, while management of mild to moderate cases is guided by clinical assessment and the identification of risk factors for complications. Dutch primary care guidelines stratify patients with acute cough into three groups: patients without pneumonia and without risk factors, those without pneumonia but with risk factors of a complicated course (age >75 years or comorbidities), and those with a clinical diagnosis of pneumonia (based on symptoms such as prolonged fever, abnormal auscultation, recent pneumonia-related hospitalisation, or CRP >100 mg/L). Dutch GPs are more likely to closely monitor and prioritise antibiotic treatment in the last two groups^13^. However, the risk factors used for stratification are largely based on expert consensus rather than robust evidence^13^.

Accurate identification of patients at risk for a complicated course of LRTIs is essential for optimising management and reducing healthcare burden. A recent systematic review by Rijk et al.^14^ identified several promising prognostic factors for complicated LRTI, including increasing age, male sex, current smoking, diabetes, history of stroke, cancer, heart failure, hospitalisation in the previous year, the absence of influenza vaccination, current use of systemic corticosteroids, antibiotic use in the previous month, a respiratory rate > 25/min, and a diagnosis of pneumonia^14^. However, the quality of evidence for these prognostic factors is limited^14^, and as these factors have not yet been combined into a validated prediction model for use in primary care, their independent predictive value remains uncertain.

Additionally, studies from the United States, United Kingdom, and the Netherlands link low socioeconomic status (SES), area-deprivation, and ethnic minorities to an increased incidence of a complicated course of LRTIs^15-21^. Nevertheless, SES and migration background have not yet been evaluated as potential predictors of complicated LRTIs, nor are they considered in current clinical guidelines, which may contribute to persisting health inequalities^13^.

In this study, we aim to evaluate the added value of SES and migration background as predictive factors of a complicated course of LRTIs in primary care. By integrating these factors with established conventional prognostic factors included in current guidelines, we aim to generate evidence to support future improvements in risk stratification and promote more equitable, data-informed care.

## Methods

### Study Design

We conducted a retrospective cohort study using routinely collected primary care data from general practices participating in the Extramural LUMC Academic Network (ELAN)^41,42^, covering the period between January 1, 2014, and December 31, 2023, excluding the COVID-19 period (January 1, 2020 – March 31, 2022), to minimize pandemic-related bias. The ELAN-GP database^41,42^ comprises electronic health records from approximately 750,000 patients registered with affiliated general practitioners in the regions of The Hague, Zoetermeer, and Leiden. These pseudonymised records were linked at the individual level to sociodemographic and hospital insurance claims data from Statistics Netherlands (SN) within the SN secure remote access environment^41,42^. Data from 2014 to 2022 were used to develop the prediction models and internal validation (development cohort), and data from 2023 were used for temporal external validation (validation cohort).

### Participants

Patients were included if they were aged 18 years or older and presented to their GP with a new episode of an uncomplicated LRTI during the study period. LRTI episodes were identified using the following International Classification of Primary Care (ICPC) codes: acute bronchitis/bronchiolitis (R78), bronchiolitis (R78.01), influenza (R80), pneumonia (R81), SARS-CoV-2 (R83.03), and acute coughing (R05). Patients were excluded if they: 1) were hospitalised or died on the same day as the GP visit, as these episodes were already considered complicated; 2) had a new diagnosis of a major pulmonary condition (lung and respiratory tract malignancies, chronic obstructive pulmonary disease (COPD), Asthma, cystic fibrosis, pulmonary embolism) within 30 days following the GP consultation, to avoid misclassification of other acute respiratory conditions as LRTI; 3) had a prior LRTI-related GP visit within six weeks, to exclude persistent symptoms; 4) were nursing home residents at the time of the LRTI episode, due to their distinct frailty profiles and risk factors. Multiple LRTI episodes per patient were allowed, provided they were separated by at least six weeks. To account for within-patient correlation due to repeated episodes, robust standard errors were calculated accounting for clustering at the patient level.

The ICPC-codes used to define major lung conditions are provided in Supplementary table M1

### Ethics

The appropriate approval that the study was not subject to the Medical Examination Act was granted by the Medical Ethical Committee of the Leiden University Medical Center (LUMC) under reference number 25-3023.

### Outcome measure

The primary outcome was a complicated course of LRTI, defined as all-cause hospitalisation or death within 30 days of the index GP consultation.

### Predictor variables

Predictor selection was based on a prior systematic review of Rijk et al.^14^, using the factors identified as promising prognostic factors. Of these, the following variables were selected for our analyses: age at consultation, sex, current smoking, diabetes mellitus, a history of stroke, cancer, or heart failure, hospitalisation in the previous year, current use of systemic corticosteroids, antibiotic prescription in the previous month before consultation, and clinical diagnosis of pneumonia at consultation. Although a respiratory rate ≥25/min and having received an influenza vaccination in the previous year were also identified as promising prognostic factors in the review, these factors could not be included in the present study as data were not available in the SN and ELAN databases. Based on clinical relevance and prior findings, additional variables were considered: same-day antibiotic prescriptions and same-day corticosteroid prescriptions in those with pre-existing asthma or COPD (associated with risk profile, disease severity, and course of disease), and pre-existing pulmonary disease (asthma, COPD, or cystic fibrosis). In practices using point-of-care CRP testing, CRP results were also included as a sub analysis, as these guide treatment decisions and reflect disease severity.

Current smoking was defined as a binary variable (yes/no), based on the status recorded closest to the GP consultation. Smoking status was derived using structured fields and unstructured free-text through text mining by using string matching^43^. Patients classified as ‘former smoker’ or missing were assumed to be non-current smokers. This assumption was validated by comparing overall and subgroup-specific smoking prevalence in our cohort to national statistics, which showed similar distributions.

Comorbidities were identified through Diagnosis Treatment Combination (DTC) codes (hospital care) and through ICPC-codes (primary care). A comorbidity was considered present if registered in either setting. Current corticosteroid use was defined as having an active prescription by the GP on the consultation date, excluding those initiated on the same day. Missing prescription end dates (4.6%) were imputed through multiple imputation by chained equations (MICE), and averaged across the imputations. Furthermore, antibiotic use in the previous month was defined as having a prior prescription by the GP within 30 days before the index consultation. A clinical diagnosis of pneumonia was defined as a GP diagnosis of pneumonia (ICPC R81) recorded on the day of consultation. Point-of-care CRP test results >100 mg/L were highly correlated with clinical pneumonia diagnosis. Therefore, in the CRP subgroup analysis, both variables were combined and categorised as: 1) CRP not measured/ unknown; 2) CRP 0-20 mg/L; 3) CRP 20-100 mg/L; 4) CRP >100 mg/l or clinical diagnosis of pneumonia.

SES was defined using household-level financial prosperity, using a composite measure of the standardised annual disposable income and wealth according to the definition of SN, which categorises this composite measure into percentiles within the Dutch population^44^. SES was categorised into five groups according to quintiles in the Dutch population. Migration background was determined by an individual’s country of birth when born abroad or the parent’s country of birth when born in the Netherlands, in accordance with the SN definition^44^.

The DTC-, ATC– and ICPC-codes regarding diagnoses and medication are listed in Supplementary Tables M1 and M2.

### Statistical Analyses

Population characteristics were described using frequencies and percentages or medians with interquartile range (IQR). All variables were classified as dichotomous except for age, SES (ordinal), and migration background (nominal). Multivariable logistic regression models with patient-level clustering were used to develop the prediction models.

#### Development of prediction models

Data from January 1, 2014, until December 31, 2022, excluding the COVID-19 period (January 1, 2020 to March 31, 2022), were used to develop three different prediction models: 1) ‘the conventional model’, including all prespecified risk factors minus SES and migration background; 2) ‘the SES model’, which added SES categories to the conventional model; 3) ‘the migration model’, which additionally included migration background. Multicollinearity was assessed using Pearson correlation coefficients; values > 0.8 indicates were considered indicative of high collinearity, but none were detected. Interaction terms were tested between key variables (age, sex, SES, migration background, and same-day antibiotic prescription) by subsequently adding interactions in multivariable logistic regression models with all prespecified risk factors. Interactions which yielded a p-value <0.05 and increased the model fit based on likelihood ratio tests were found between same-day antibiotic prescription and pneumonia diagnosis, and between diabetes mellitus and age. Least Absolute Shrinkage and Selection Operator (LASSO) selection procedures were performed separately on the three different multivariable logistic regression models with the prespecified candidate predictors and interaction terms to select the predictive variables in each model as well as to prevent overfitting and enhance model interpretability. After the selection of predictive variables by the LASSO method, multivariable logistic regression models with the selected variables were used to develop the final predictive models to yield coefficients and probabilities. Model performance was assessed by goodness-of-fit, Brier-score, discrimination and calibration. Goodness of fit was compared between models using the Akaike Information Criterion (AIC), with differences > 10 considered to provide strong support for the better-fitting model^23^. The discriminative abilities of the models were assessed using the area under the receiving operator curve (AUROC), which corresponds to the C-statistic. Calibration was evaluated and compared through the slope and intercept of calibration plots.

Observations with missing values were omitted from analyses if data on one of the included variables was missing (0.6% of cases), except for smoking status which was handled pragmatically by imputing missing values as non-current smokers, and corticosteroid use which was imputed as described above. Odds ratio’s and their corresponding 95% confidence intervals were derived from the multivariable logistic regression models with p-values <0.05 considered significant.

We calculated observed absolute risks of a complicated LRTI course in the development cohort, stratified by the Dutch primary care clinical guideline (NHG) risk groups^13f^ and SES categories. In parallel, we used the SES model to estimate the increase in mean predicted probability across the same strata to evaluate alignment between observed outcomes and model-based risk. By plotting absolute risks, we aim to enhance the clinical interpretability and relevance of found socio-economic disparities within familiar clinical categories.

Sub-analyses were performed for practices using point-of-care CRP testing, replacing the clinical pneumonia diagnosis with the combined CRP/pneumonia classification. All analyses were performed in Stata (Stata Corp. 2023. Stata Statistical Software: Release 18. College Station, TX: Stata Corp LLC.).

#### Internal validation

The three models were internally validated by bootstrapping with 1,000 iterations to estimate model stability. The discriminative ability, calibration and goodness of fit were subsequently compared between models.

#### Temporal validation

Temporal validation was performed using data from the 2023 cohort. AUROC values of the models were compared to those in the development cohort.

## Results

### Population characteristics

The development cohort (2014-2022 without the COVID-19 period) comprised 186,094 LRTI episodes from 145,445 patients. Of these, 4,072 episodes (2.19%) progressed to a complicated LRTI. Demographic and clinical characteristics of the cohort, including its univariable association with a complicated course, are presented in Table 1.

**Table 1:**
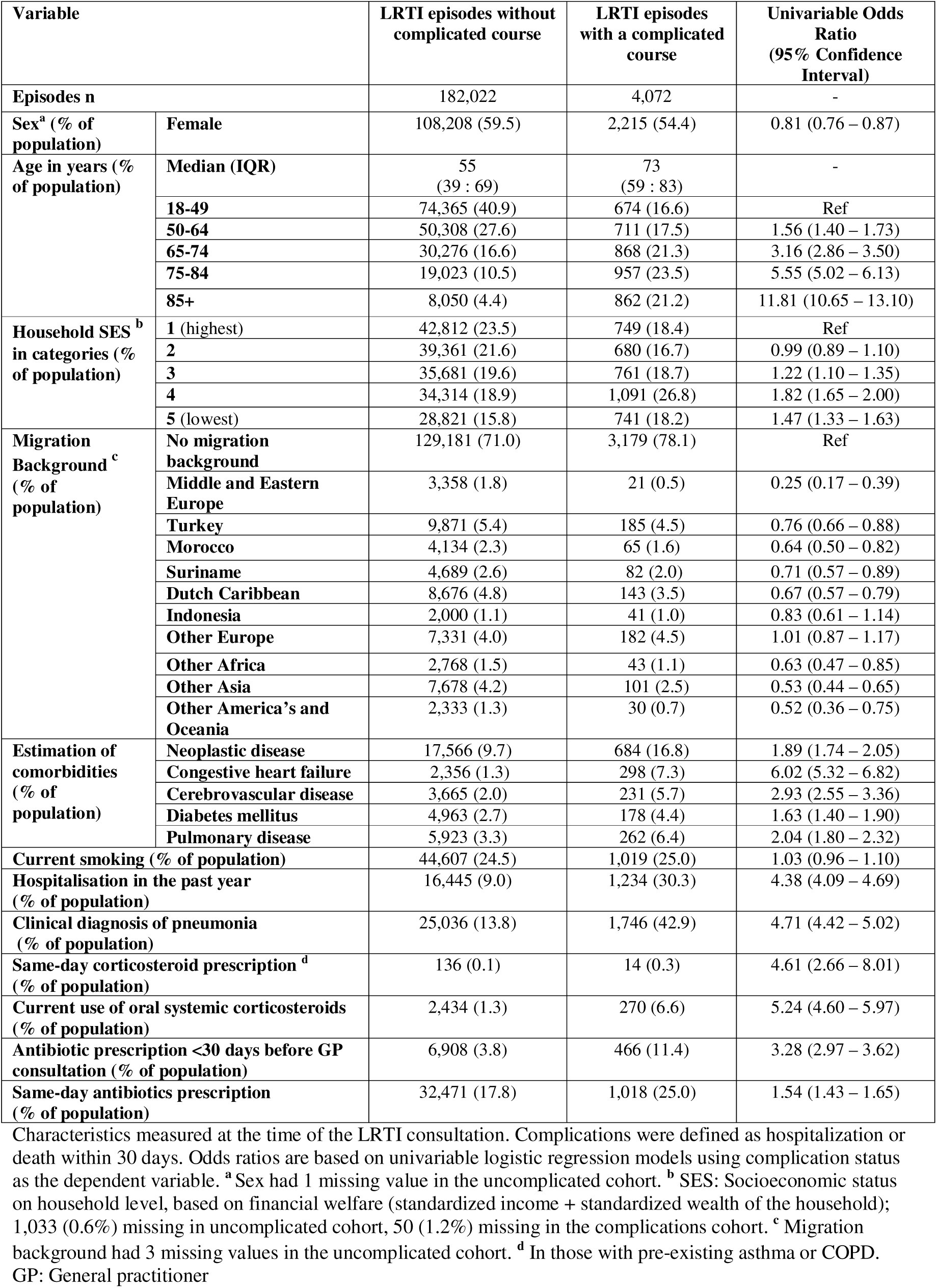
Population characteristics of the development cohort (2014-2022) and univariable associations with a complicated course of LRTI.

The median patient age was 56 years (IQR: 39 – 69) and 59.3% were female. Diabetes mellitus was present in 2.7% of the cohort, while 1.4% had a history heart failure, 9.2% had neoplastic disease, and 2.1% had a history of stroke. All comorbidities were more prevalent in those who developed a complicated LRTI (Table 1). Current smoking was recorded for 24.5% of patients, and antibiotics were prescribed on the same day for 18.0% of LRTI episodes. A clinical diagnosis of pneumonia was recorded in 14.4% of cases.

Patients in the lowest (fifth) and highest (first) SES-categories comprised 15.9% and 23.4% of the cohort, respectively. Patients in the fourth and fifth categories were overrepresented among those who developed a complicated LRTI (Table 1). A migration background was recorded in 28.9% of patients; the most common were ‘other Europe’ (5.4%), Suriname (4.7%), ‘other Asia’ (4.2%), and Indonesia (4.0%), while 71.1% had no migration background.

The temporal validation cohort (2023) included 25,756 LRTI episodes, of which 585 (2.27%) progressed to a complicated LRTI. Characteristics of this cohort are detailed in Supplementary Table S2.

### Development of prediction models

Three multivariable logistic regression models were developed: 1) ‘the conventional model’, including all prespecified risk factors minus SES and migration background; 2) ‘the SES model’, which added SES categories to the conventional model; 3) ‘the migration model’, which additionally included migration background.

In the conventional model, the strongest predictors for complicated LRTI were increasing age, with an adjusted Odds Ratio (aOR) of 6.43 (95%CI: 5.71 – 7.23) for > 85 years, pneumonia diagnosis during GP consultation (aOR 3.10 [95%CI: 2.83 – 3.39]), current use of oral systemic corticosteroids (aOR 2.10 [95%CI: 1.81 – 2.44], hospitalisation in the past year (aOR 1.96 [95%CI: 1.80 – 2.13]), and diabetes mellitus (aOR 1.95 [95%CI: 1.21 – 3.16]) (Table 2). In the SES model, lower SES was an independent predictive factor for complicated LRTI, with the lowest SES-category demonstrating the highest odds (aOR 1.46 [95%CI: 1.31 – 1.62]) compared to the highest category (Table 2). In the migration model, those with a Middle and Eastern European background had an aOR of 0.55 (95%CI: 0.35 – 0.85) compared to those without a migration background, while other migration backgrounds showed no statistically significant associations (Table 2). Effect estimates for conventional risk factors remained stable across the three models (Table 2).

**Table 2:** Multivariable associations for a complicated course of LRTI from the conventional, SES, and migration prediction model.

|  | Conventional model<br>aOR (95% CI) | SES model<br>aOR (95% CI) | Migration model<br>aOR (95% CI) |
| --- | --- | --- | --- |
| **Sex** |  |  |  |
| Male | Ref | Ref | Ref |
| Female | 0.90 (0.84 – 0.96) | 0.88 (0.82 – 0.94) | 0.88 (0.83 – 0.94) |
| **Age** |  |  |  |
| 18-49 | Ref | Ref | Ref |
| 50-64 | 1.35 (1.21 – 1.50) | 1.41 (1.27 – 1.58) | 1.39 (1.25 – 1.55) |
| 65-74 | 2.36 (2.12 – 2.63) | 2.45 (2.20 – 2.72) | 2.41 (2.16 – 2.68) |
| 75-84 | 3.50 (3.14 – 3.90) | 3.50 (3.14 – 3.91) | 3.46 (3.09 – 3.86) |
| >85 | 6.43 (5.71 – 7.23) | 6.36 (5.65 – 7.16) | 6.25 (5.54 – 7.05) |
| **Household SES in categories** |  |  |  |
| 1 (highest) | X | Ref | Ref |
| 2 | X | 1.11 (1.00 – 1.24) | 1.11 (1.00 – 1.23) |
| 3 | X | 1.27 (1.14 – 1.41) | 1.27 (1.14 – 1.41) |
| 4 | X | 1.36 (1.23 – 1.50) | 1.36 (1.23 – 1.51) |
| 5 (lowest) | X | 1.46 (1.31 – 1.62) | 1.47 (1.31 – 1.64) |
| **Migration Background** |  |  |  |
| The Netherlands | X | X | Ref |
| Middle & Eastern Europe | X | X | 0.55 (0.35 – 0.85) |
| Other Europe | X | X | 0.88 (0.75 – 1.03) |
| Turkey | X | X | 1.02 (0.78 – 1.32) |
| Morocco | X | X | 0.96 (0.76 – 1.21) |
| Suriname | X | X | 0.99 (0.83 – 1.17) |
| Dutch Caribbean | X | X | 1.24 (0.90 – 1.71) |
| Indonesia | X | X | 1.09 (0.93 – 1.27) |
| Other Africa | X | X | 1.15 (0.84 – 1.59) |
| Other Asia | X | X | 0.91 (0.74 – 1.13) |
| Other America's and Oceania | X | X | 1.02 (0.71 – 1.46) |
| **Comorbidities** |  |  |  |
| Neoplastic disease | 1.68 (1.54 – 1.84) | 1.70 (1.56 – 1.86) | 1.70 (1.55 – 1.86) |
| Congestive heart failure | 1.73 (1.50 – 2.00) | 1.71 (1.48 – 1.98) | 1.72 (1.49 – 1.98) |
| Cerebrovascular disease | 1.21 (1.04 – 1.40) | 1.20 (1.03 – 1.39) | 1.20 (1.03 – 1.39) |
| Diabetes mellitus | 1.95 (1.21 – 3.16) | 1.89 (1.17 – 3.05) | 1.87 (1.15 – 3.02) |
| Pulmonary disease | 1.20 (1.04 – 1.38) | 1.17 (1.02 – 1.36) | 1.17 (1.02 – 1.36) |
| **Health** |  |  |  |
| Current smoking | 1.08 (1.00 – 1.17) | 1.05 (0.97 – 1.13) | 1.05 (0.97 – 1.13) |
| Hospitalisation in the past year | 1.96 (1.80 – 2.13) | 1.92 (1.77 – 2.09) | 1.92 (1.77 – 2.09) |
| Current use of oral systemic corticosteroids | 2.10 (1.81 – 2.44) | 2.09 (1.80 – 2.42) | 2.09 (1.80 – 2.42) |
| Antibiotics prescribed <30 days before GP consultation | 1.38 (1.22 – 1.55) | 1.37 (1.22 – 1.54) | 1.37 (1.22 – 1.54) |
| **Degree of illness** |  |  |  |
| Clinical diagnosis of pneumonia | 3.10 (2.83 – 3.39) | 3.09 (2.83 – 3.38) | 3.09 (2.83 – 3.38) |
| Same-day antibiotics prescription | 1.09 (0.97 – 1.23) | 1.09 (0.97 – 1.22) | 1.09 (0.97 – 1.22) |
| Same-day corticosteroid prescription <sup>a</sup> | 1.42 (0.75 – 2.70) | 1.44 (0.76 – 2.72) | 1.45 (0.77 – 2.73) |
| **Interactions** |  |  |  |
| Same-day antibiotics*Pneumonia | 0.80 (0.68 – 0.94) | 0.81 (0.69 – 0.95) | 0.81 (0.689 – 0.95) |

| diagnosis |  |  |  |
| --- | --- | --- | --- |
| Age category 18-49*<br>Diabetes Mellitus | Ref | Ref | Ref |
| Age category 50-64*<br>Diabetes Mellitus | 0.92 (0.52 – 1.61) | 0.90 (0.51 – 1.58) | 0.91 (0.52 – 1.60) |
| Age category 65-74*<br>Diabetes Mellitus | 0.63 (0.35 – 1.11) | 0.62 (0.35 – 1.09) | 0.62 (0.35 – 1.10) |
| Age category 75-84*<br>Diabetes Mellitus | 0.55 (0.31 – 0.99) | 0.55 (0.31 – 0.99) | 0.56 (0.31 – 1.00) |
| Age category >85*<br>Diabetes Mellitus | 0.35 (0.18 – 0.69) | 0.36 (0.18 – 0.71) | 0.37 (0.19 – 0.72) |

Adjusted odds ratios are derived from the multivariable logistic regression models. Variables in all final models were selected through LASSO selection procedures, which did not exclude any prespecified variables. Patients with missing variables were excluded from analyses (n=1,087, 0.6%). <sup>a</sup> In those with pre-existing asthma or COPD. CI: Confidence interval. aOR: adjusted Odds Ratio. GP: General practitioner

### Comparison of performance, calibration, and fit of prediction models

All models demonstrated acceptable-to-good discriminative ability with cross-validated AUROCs of 0.78 (95%CI: 0.77 – 0.79) for the conventional model, 0.78 (95%CI: 0.77 – 0.79) for the SES model, and 0.78 (95%CI: 0.77 – 0.79) for the migration model (table 3). Each model also showed similar Brier-scores (0.020) and good calibration, indicated by calibration intercepts close to 0 and slopes near 1.0 (Table 3), consistent with minimal overfitting^22^. Calibration plots for the three models are provided in the supplementary materials figures F1, F2, and F3. Despite similar discrimination and calibration, the migration model showed the best overall fit, as indicated by the lowest Akaike information criterion (AIC) value (33966.76), compared to the SES model (33980.46) and the conventional model (34045.55) (Table 3). This suggests that, among the three, the migration model most effectively explains variation in the observed data^23^. However, the largest improvement in model fit occurred with the addition of socioeconomic status; the incremental gain from including migration background was comparatively modest.

**Table 3.** Model performance metrics for the conventional model, SES model and migration model for the development dataset.

|  | Conventional model | SES model | Migration model |
| --- | --- | --- | --- |
| Cross-validated AUROC (95%CI) | 0.78 (0.77 – 0.79) | 0.78 (0.77 – 0.79) | 0.78 (0.77 – 0.79) |
| AUROC bootstrap sample (95%CI) | 0.77 (0.77 – 0.78) | 0.78 (0.77 – 0.79) | 0.78 (0.78 – 0.79) |
| Brier-score | 0.020 | 0.020 | 0.020 |
| Calibration intercept (95% CI) | 0.01 (-0.08 – 0.11) | 0.01 (-0.08 – 0.11) | 0.04 (-0.06 – 0.13) |
| Calibration slope (95%CI) | 1.004 (0.977 – 1.031) | 1.004 (0.977 – 1.031) | 1.012 (0.985 – 1.039) |
| AIC* | 34045.55 | 33980.46 | 33966.76 |

\*Akaike information criterion. CI: Confidence interval.

Internally validating the models by bootstrapping with 1,000 iterations yielded similar discrimination, calibration and goodness-of-fit statistics.

### Risk assessment

To evaluate how model-predicted risk aligns with clinical guideline categories, we visualised both observed and predicted risks of a complicated course of LRTI across Dutch general practitioner society (NHG)-defined strata^13^: 1) patients without pneumonia at time of LRTI consultation and without risk factors; 2) patients without pneumonia at time of LRTI consultation but with risk factors; 3) patients with a clinical diagnosis of pneumonia at time of LRTI consultation.

In all NHG-defined risk strata, a gradient was observed in both observed and predicted risks across SES categories. Patients in lower SES categories consistently had higher absolute and predicted risks of a complicated course of LRTI. The difference was most pronounced in the pneumonia group, where absolute risk was 7.2% in the lowest SES category compared to 5.0% in the highest, while the predicted risk was 7.5% in the lowest SES category compared to 4.9% in the highest (Figure 1 and Supplementary Table S4). Additionally, in the pneumonia group, the mean predicted probability of a complicated course of LRTI was 2.1% higher in the lowest SES category compared to the highest (Figure 2).

**Figure 1:**
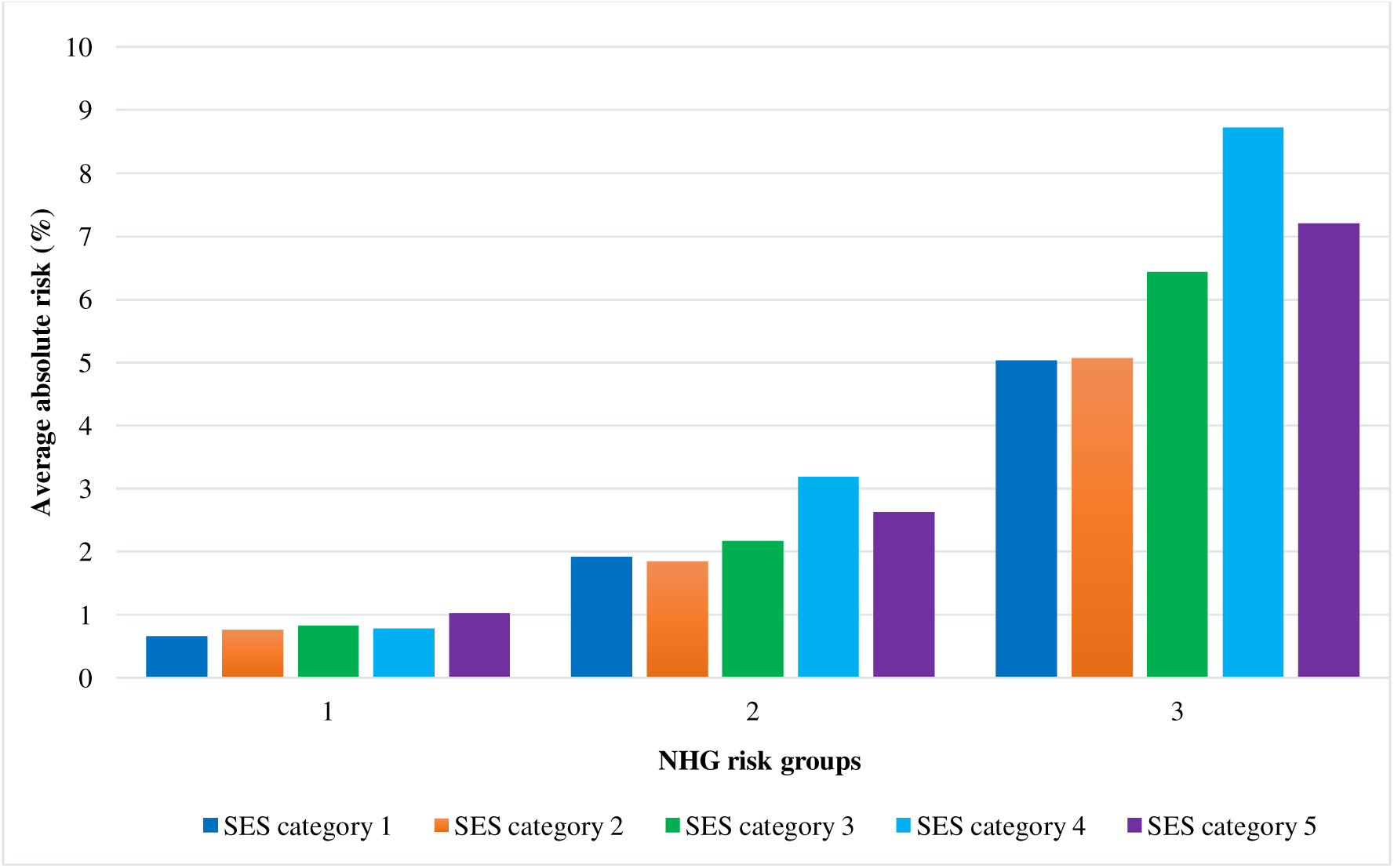
Observed absolute risk of a complicated course of LRTI across NHG guideline risk strata SES: socioeconomic status. NHG: Dutch general practitioner society. Absolute average risk was estimated in the development dataset by SES category across NHG guideline risk strata. Risk strata 1: those without risk factors or pneumonia. Risk strata 2: those with risk factors: age >75 years, heart or lung disease, diabetes mellitus, neurological disease, liver or kidney disease, immunocompromised. Risk strata 3: those with a clinical diagnosis of pneumonia which is based on clinical presentation, recent pneumonia-associated hospitalisation, or point-of-care CRP >100 mg/L.

**Figure 2:**
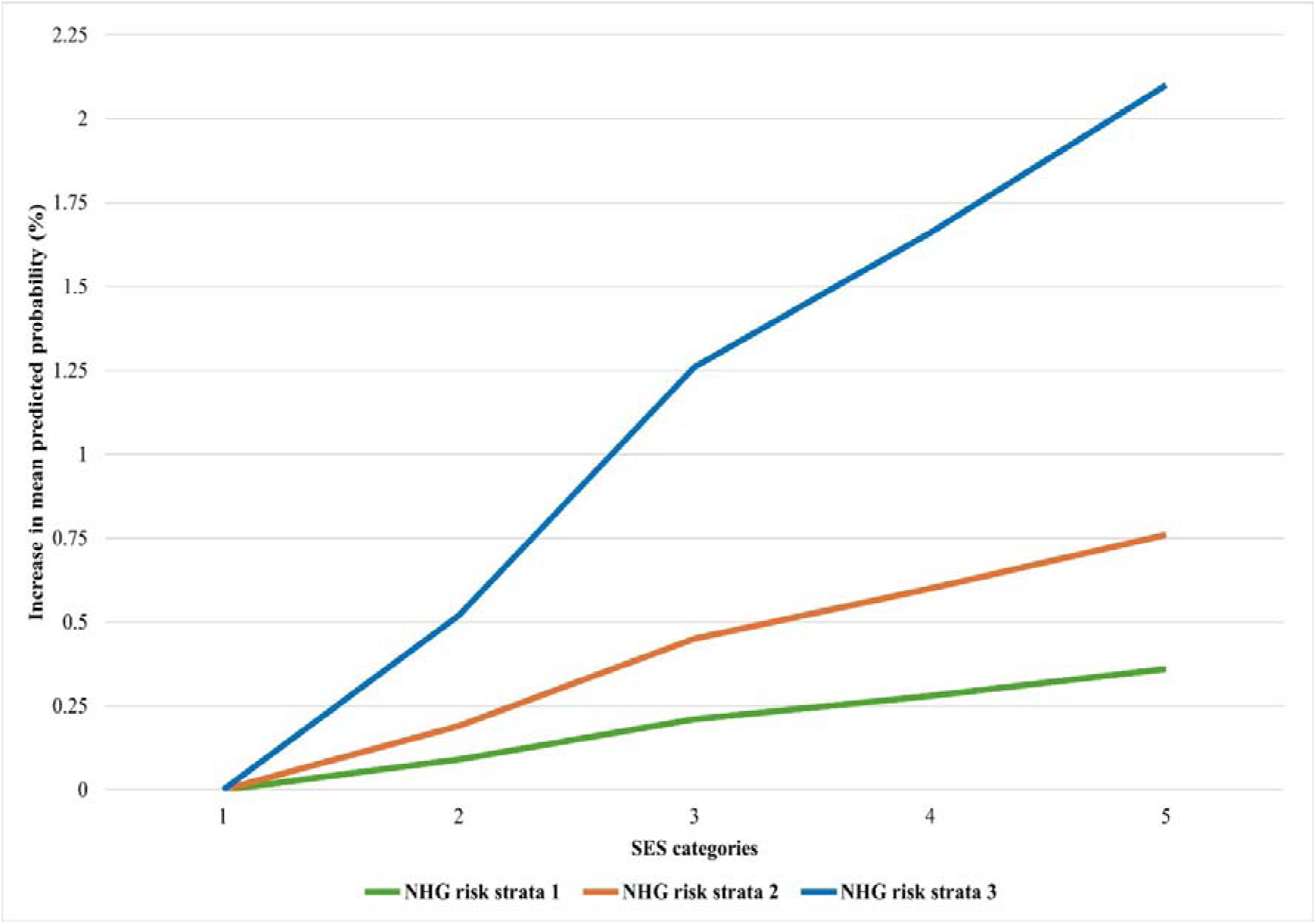
Increase in mean predicted probability of a complicated course of LRTI compared to SES category 1 across NHG risk strata, adjusted for all model covariates SES: Socioeconomic status. NHG: Dutch general practitioner society. Differences in predicted risk of a complicated course of LRTIs of SES categories compared to the first category in the development cohort where five marks the lowest SES. The SES model was used for analyses. The 95% confidence intervals are provided in Supplementary Table S6 as boundaries were too small to visually depict. Risk strata 1: those without risk factors or pneumonia. Risk strata 2: those without pneumonia with risk factors: age >75 years, heart or lung disease, diabetes mellitus, neurological disease, liver or kidney disease, immunocompromised. Risk strata 3: those with a clinical diagnosis of pneumonia which is based on clinical presentation, recent pneumonia-associated hospitalisation, or point-of-care CRP >100 mg/L.

### Temporal validation

All models showed acceptable discrimination in the 2023 validation cohort, with AUROCs slightly higher than those in the development cohort (Table 4). Differences in discrimination, calibration, and goodness of fit were similar to those observed in the development cohort.

**Table 4.** Model performance metrics for the conventional model, SES model and migration model for the validation dataset.

|  | Conventional model | SES model | Migration model |
| --- | --- | --- | --- |
| Cross-validated AUROC (95%CI) | 0.78 (0.77 – 0.81) | 0.79 (0.77 – 0.81) | 0.79 (0.77 – 0.81) |
| AUROC bootstrap sample (95%CI) | 0.77 (0.76 – 0.80) | 0.79 (0.76 – 0.81) | 0.79 (0.77 – 0.81) |
| Brier-score | 0.021 | 0.021 | 0.021 |
| Calibration intercept (95% CI) | -0.04 (-0.29 – 0.20) | -0.06 (-0.30 – 0.19) | -0.03 (-0.28 – 0.22) |
| Calibration slope (95%CI) | 1.005 (0.932 – 1.077) | 1.002 (0.930 – 1.074) | 1.010 (0.937 – 1.082) |
| AIC* | 4882.62 | 4872.57 | 4871.49 |

\*Akaike information criterion. CI: Confidence interval.

### Sub-analyses of patients with C-reactive protein testing

Among the 156 GP practices included in the database, 124 implemented point-of-care CRP-testing in patients presenting with LRTI, before 2023. Subgroup analyses were conducted from the moment the practice began using CRP testing. In this subgroup, CRP values were found to strongly correlate with a clinical diagnosis of pneumonia. Based on this, we defined a four-category variable combining CRP results and pneumonia diagnosis: 1) CRP not measured/unknown; 2) CRP 0-20 mg/L; 3) CRP 20-100 mg/L; 4) CRP >100 mg/l or clinical diagnosis of pneumonia. The last category showed a significantly increased adjusted odds ratio of a complicated LRTI course (aOR 3.24, 95% CI: 1.66 – 6.32) compared to a CRP 0-20, consistent with findings from the main model using clinical pneumonia diagnosis alone. Associations between other covariates and complicated LRTI remained stable in this subgroup analysis and are presented in Supplementary Tables S7-S10.

## Discussion

In this large retrospective primary care cohort, we found that lower SES was an independent predictor of a complicated course of LRTI in adults, even after adjusting for conventional clinical factors such as age, comorbidities and smoking. Incorporating SES into a model with conventional risk factors significantly improved the model fit, while migration background added only marginal explanatory value. Notably, we observed a consistent SES gradient in both absolute and predicted risks across existing clinical guideline strata, suggesting that SES captures additional vulnerability not currently accounted for in primary care risk stratification.

Our findings extend previous research on LRTI prediction models^24-27^, by integrating SES and migration background into the analyses. Notably, lower SES retained its predictive value after adjusting for conventional risk factors. These results underscore that SES may serve as a practical proxy for a range of underlying risk factors relevant to infection risk and the course of disease. These may include housing quality, air pollution, vaccination uptake, working conditions, health literacy, psychological factors, access to care, and lifestyle factors such as obesity, alcohol consumption, and smoking^28-34^; most of which are not captured during GP consultations.

Although previous studies have reported associations between socioeconomic factors and complicated LRTI^15-19^, most used population-level data. By contrast, our study begins at the point of GP consultation, providing more detailed insight into individual disease trajectories. We observed a clear SES gradient in both observed and predicted risk across clinical guideline-defined risk strata, particularly among patients with a clinical diagnosis by the GP at first consultation of pneumonia. Within this group, predicted risk of complication ranged from 4.9% on average in the highest SES category to 7.5% on average in the lowest, while the mean predicted probability of a complicated course of LRTI increased with 2.1% from the highest to the lowest category; a clinically relevant finding. Using SES-informed stratification therefore enables us to identify vulnerable patients currently missed by guideline-based risk groups.

Migration background, in contrast, showed limited predictive utility after adjustment for SES and other factors. Individuals with Middle or Eastern European backgrounds had lower adjusted odds of developing complications compared to those without a migration background. Immigrants from Middle and Eastern European countries are typically relatively healthy seasonal workers in physically demanding jobs and are, on average, younger than 35 years^35,36^. Additionally, their stay in the Netherlands is often short-term, usually less than five years^37^. These factors may contribute to the “healthy immigrant effect”, a phenomenon in which younger migrant populations, often migrant workers, tend to arrive in better overall health than the native population^38^. However, this population of migrant workers is also known to experience reduced access to healthcare services^36,39^. This combination of better baseline health and healthcare access barriers may possibly contribute to lower hospitalisation rates. This should not be interpreted as evidence that these individuals require less clinical attention; rather, it may reflect under-recognition of complications due to access barriers. As such, we do not recommend modifying clinical strategies based on these findings.

In contrast to our findings, studies from the United States linked ethnic minorities to influenza-associated hospitalisation and mortality incidence^20,21^. The discrepancy in these findings may partly be explained by major differences in healthcare systems, such as the absence or presence of universal coverage and mandatory basic insurance. Furthermore, studies on the effect of ethnicity and migration background not accounting for SES may have overlooked the confounding effect of SES and attributed mostly SES-driven associations to ethnicity or migration background. Our findings underscore the importance of separating structural determinants such as SES from ethnic or migration-based categories when developing prediction models.

Our model performed well in internal and temporal validation, achieving higher discrimination (AUROC/C-statistic 0.78) than previously published primary care prediction models for LRTIs (with C-statistics ranging from 0.63 to 0.74)^24,25,27^. While we lacked some clinical metrics such as oxygen saturation, blood pressure, and respiratory rate^25,27^, we accounted for disease severity through proxies including pneumonia diagnosis, CRP results in our sub analysis, and medication prescription patterns. The current quality of evidence supporting many of the previously identified promising prognostic factors has been low to very low^14^. The relative robustness of conventional risk factors for complicated LRTIs in our models supports and extends earlier evidence^14,24-26,40^, while demonstrating the added value of incorporating SES.

In 2020, only 0.2% of the Dutch population reported unmet needs for medical care, underscoring that access to care is generally not a significant barrier within the Dutch healthcare system^4^. Nevertheless, our findings demonstrate that health inequalities persist in the context of complicated LRTIs, similar to other, health outcomes^5,6,8,9^. This reinforces the notion that disparities are driven not solely by access barriers, but also by broader social determinants. Given the similar population composition and healthcare infrastructure of many European countries, our results likely generalise beyond the Dutch context. Indeed, similar patterns of social deprivation and poor LRTI outcomes have been reported in the United Kingdom (UK)^18^. However, few prior studies have examined SES and migration background jointly.

Strengths of our study include its large sample size, representative primary care setting, and linkage of real-world clinical data to detailed sociodemographic data and hospital outcomes. The gatekeeper role of Dutch GPs allowed for unique insights into LRTI course of disease from first presentation. Moreover, by including conventional risk factors, SES, and migration background allowed for a more nuanced understanding of their relative contributions. Limitations include the observational nature of the study, precluding causal inferences. Influenza vaccination status was unavailable, which may be one intermediate factor linking SES with LRTI complications. However, given that our study included all LRTIs, of which influenza represents only a subset, the impact of this limitation on our findings is likely limited. Smoking status was handled pragmatically by imputing missing values as non-current smokers; while the percentage of smokers was similar to Dutch and regional averages, some misclassification may have been possible. Furthermore, as corticosteroid and antibiotic prescription dates were only available in the GP data, some underreporting might be possible. Additionally, as we were only able to internally and temporally validate the prediction models it is uncertain how these models would perform in other parts of the Netherlands or in other countries. Finally, the model was designed to estimate risk under current care rather than directly guide clinical decision-making; further work is needed to translate it into actionable tools.

Our findings strongly support integrating SES into future prediction models and primary care guidelines for LRTI. Doing so would likely improve the early identification of high-risk individuals, and enable more targeted interventions, such as follow-up consultations or treatment and referral decisions. Furthermore, our findings might support the design of tailored public health strategies, such as focused vaccination campaigns in low-SES neighbourhoods where uptake is typically lower. Leveraging routinely available indicators such as postcode-based deprivation indices and integration within electronic health records could facilitate this integration in practice. To operationalise SES in clinical workflows, future research is needed on feasible implementation strategies and their impact on patient outcomes.

In conclusion, our study establishes SES as a significant and independent predictor of a complicated course of LRTIs, even after accounting for conventional risk factors. Our findings provide a foundation for more socially responsive risk stratification, offering a concrete step toward recognizing and addressing SES-linked vulnerability in patients with LRTIs.

## Data availability

The dataset cannot be shared publicly due to the current Dutch legislation for data protection of Statistics Netherlands data. Under certain conditions, the dataset and additional microdata are accessible for statistical and scientific research and must be directly requested from Statistics Netherlands.

## Code availability

The Stata scripts used will be disclosed through a repository on GitHub: https://github.com/elan-dcc/VanDokkum25.

## Contributions

All authors contributed equally to the conceptualisation of the study. Study design and methodology: HB CvN EDvD SlC. Literature search: EDvD and NK with contributions from HB and CvN. Data curation and data analysis: EDvD and NK with substantial contributions from HB and SlC. Data interpretation: EDvD, NK, HB and CvN with substantial contributions from all authors. Visualisation of results: EDvD, NK, HB and CvN with contributions of all authors. EDvD wrote the first draft of the manuscript with inputs from NK, HB, and CvN. All authors reviewed and revised drafts of the manuscript and approved the final version. EDvD, NK, and HB have full access to and have verified all the study data provided for the analysis.

