## Supplementary table M1 for "The predictive value of socioeconomic status and migration background for complicated lower respiratory tract infections in primary care"

**M-Tables**

**Table M1** – DTC and ICPC-codes used for definition of variables and LRTIs

**Table M2** – Drugs by ATC4-codes, used to help define Diabetes Mellitus, Immunosuppression, Antibiotics and corticosteroids.

**S-Tables**

**Table S1** – Population characteristics stratified by antibiotic prescription on the same day

**Table S2** – Population characteristics of the validation dataset

**Table S3** – Difference in deviance and deviance ration between models

**Table S4** – Absolute risk and predicted risk of a patient’s risk of developing a complicated LRTI classified into risk-groups according to NHG guidelines, stratified by SES-quintiles in derivation dataset

**Table S5** – Absolute risk and predicted risk of a patient’s risk of developing a complicated LRTI classified into risk-groups according to NHG guidelines stratified by SES-quintiles in validation dataset

**Table S6** – Increase in mean predicted probability of a complicated course of LRTI across NHG risk strata, adjusted for all model covariates

**Table S7** – Multivariable logistic regression sub analyses with CRP added in the model, only practices where a CRP was possible are included

**Table S8** – Difference in discrimination between models with CRP-POC test integrated

**Table S9** – Difference in calibration between models with CRP-POC test integrated

**Table S10** – Difference in deviance and deviance ratio between models with CRP-POC test integrated

**F-Figures**

**Figure F1 –** Calibration plot conventional model

**Figure F2 –** Calibration plot SES model

**Figure F3 –** Calibration plot migration model

**M-References**

**Table M1 – DTC- and ICPC-codes used for definition of variables and LRTI**

| **Definition** | **DTC-codes** | **ICPC-codes** |
| --- | --- | --- |
| **Lower respiratory tract infection** | **-** | R05, R78, R78.01, R80, R81, R83.03 |
| **Neoplastic disease [1]** | 0302#21, 0302#60, 0302#61, 0302#62, 0302#63, 0302#64, 0302#65, 0302#66, 0302#67, 0302#68, 0302#69, 0302#72, 0302#84, 0303#303, 0303#306, 0303#318, 0303#319, 0303#331, 0303#332, 0303#333, 0303#334, 0303#335, 0303#346, 0303#347, 0303#349, 0303#350, 0303#352, 0303#353, 0303#357, 0303#358, 0303#360, 0303#363, 0303#367, 0303#370, 0305#1110, 0305#1140, 0305#1150, 0306#10, 0306#16, 0306#20, 0306#30, 0306#40, 0306#45, 0306#48, 0306#50, 0306#60, 0306#69, 0306#70, 0306#78, 0306#84, 0306#92, 0307#M11, 0307#M12 0307#M13 0307#M14, 0307#M15, 0307#M16, 0307#M99, 0313#214, 0313#264, 0313#621, 0313#622, 0313#623, 0313#624, 0313#629, 0313#751, 0313#752, 0313#753, 0313#754, 0313#756, 0313#757, 0313#761, 0313#771, 0313#801, 0313#802, 0313#811, 0313#821, 0313#822, 0313#823, 0313#831, 0313#832, 0313#833, 0313#834, 0313#839, 0313#841, 0313#842, 0313#843, 0313#899, 0313#904, 0313#914, 0313#964, 0313#979, 0318#307, 0318#407, 0318#408, 0318#610, 0318#712, 0318#735, 0322#1303, 0322#1304, 0322#1305, 0322#1306, 0322#1308, 0330#202, 0330#203, 0330#213, 0330#223, 0330#233, 0330#242, 0330#243. | A79,  B72,  B73,  B74,  D74,  D75,  D76,  D77,  F74.01,  H75.01,  K72.01,  L71 (not L71.02),  N74,  R84,  R85,  S77 (not S77.01),  T71,  U75,  U76,  U77,  W72,  X75,  X76,  X77,  Y77,  Y78 |
| **Liver disease [1]** | 0313#463, 0313#941 0313#942, 0313#943, 0313#944, 0313#945, 0313#946, 0318#701, 0318#705, 0318#707, 0318#708, 0318#709, 0318#713, 0318#718. | D72, D97 |
| **Congestive heart failure [1]** | 0313#107, 0320#301, 0320#302, 0335#262. | K77 |
| **Cerebrovascular disease [1]** | 0313#121, 0330#1102, 0330#1111, 0330#1112, 0335#263. | K89, K90 |
| **Chronic renal disease [1]** | 0313#324, 0313#325, 0313#331, 0313#332, 0313#336, 0313#339. | U99.01 |
| **Pulmonary disease (other)** | 0303#309, 0313#601, 0322#1201, 0322#1241, 0322#1403, 0335#272. | R91, R95, R96, T99.10 |
| **Neurologic disease** | 0330#501, 0330#522, 0330#531, 0330#911, 0330#999, 0335#252. | N86, N87, N99.01, N99.02, N99.03 |
| **Diabetes Mellitus** | 0313#221, 0313#222, 0313#223, 0318#902, 0335#222 | T90 |
| **Immunocompromised** | 0303#551, 0303#553, 0303#554, 0303#555, 0303#557, 0303#559, 0303#560, 0303#561, 0303#562, 0303#563, 0313#70, 0313#72, 0313#73, 0313#74, 0313#76, 0313#78, 0313#79, 0313#81, 0313#82, 0313#83, 0318#761 0318#763 0318#764, 0318#766, 0318#767, 0318#768, 0328#2910, 0328#2920, 0328#2930, 0303#325, 0303#326, 0305#1394, 0313#501, 0313#503, 0313#512, 0313#515, 0313#521, 0313#522, 0313#523, 0313#524, 0313#525, 0313#526, 0313#527, 0313#922, 0313#923, 0318#601, 0318#602, 0324#101, 0324#102, 0324#114, 0324#201, 0324#202, 0324#301, 0324#302, 0324#304, 0324#305, 0324#306, 0324#307, 0324#311, 0324#312, 0324#313, 0324#315, 0324#316, 0324#317, 0324#318, 0324#319, 0313#461, 0313#462. | A87.02, B72, B73, B90, T99.01, B74.01  Rest was defined through medication use (such as immunosuppressives for colitis ulcerosa and rheumatoid arthritis) |
| **Other cardiovascular disease** | 0303#412 0303#418, 0303#419, 0303#420, 0313#124, 0313#133, 0320#202 0320#203, 0320#204, 0320#205, 0320#601, 0320#801, 0320#802, 0320#803, 0320#804, 0328#2220, 0328#2320, 0328#2400, 0328#2415, 0328#2425, 0328#2470, 0328#2550, 0328#2555, 0328#2560, 0328#2570, 0328#2585, 0328#2630, 0328#2635, 0328#2640, 0328#2645, 0328#2650, 0328#2655, 0328#2665, 0328#2720, 0328#2740, 0328#2770, 0328#2785, 0328#2940, 0328#3210, 0328#3310 | K74, K75, K76, K78, K84, K91, K92.01, K99.01 |
| **Major pulmonary conditions used for exclusion criteria (**lung and respiratory tract malignancies, COPD, Asthma, cystic fibrosis, pulmonary embolism**)** | - | R84, R85, R95, R96, T99.10, K93 |

**Table M2 – Drugs by ATC4-codes, used to help define Diabetes Mellitus, Immunosuppression, Antibiotics and corticosteroids.**

| **ATC4-code [2]** | **Name** |
| --- | --- |
| **Diabetes Mellitus Medication** | |
| A10A | Insulins and analogues |
| A10B | Blood glucose lowering drugs, excluding insulins |
| **Immunosuppressives** | |
| L04A | Immunosuppressive drugs |
| L01B | Antimetabolites |
| L01X | Other antineoplastic agents |
| H02A | Corticosteroids for systemic use, plain |
| **Antibiotics** | |
| J01A | Tetracyclines |
| J01B | Amphenicols |
| J01C | Beta-lactam antibacterials, penicillin’s |
| J01D | Other beta-lactam antibacterials |
| J01E | Sulphonamides and Trimethoprim |
| J01F | Macrolides, Lincosamides and Streptogramins |
| J01G | Aminoglycoside antibacterials |
| J01M | Quinolone antibacterials |
| J01R | Combination of antibacterials |
| J01X | Other antibacterials |

**Table S1: Population Characteristics stratified by antibiotics prescription on the same day**

| **Variables** | | **No antibiotic prescription** | **Antibiotic prescription** |
| --- | --- | --- | --- |
| **Subjects n** | | 152,605 | 33,489 |
| **Complicated LRTI (% of the population)** | | 3,054 (2.0) | 1,018 (3.0) |
| **Sex^1^** | **Female (%)** | 91,637 (60.0) | 18,786 (56.1) |
|  | **Male (%)** | 60,967 (40.0) | 14,703 (43.9) |
| **Age in years (% of population)** | **Median (IQR)** | 55  (38 : 68) | 59  (43 : 71) |
|  | **18-49** | 63,556 (41.6) | 11,483 (34.3) |
|  | **50-64** | 41,794 (27.4) | 9,225 (27.6) |
|  | **65-74** | 24,813 (16.3) | 6,331 (18.9) |
|  | **75-84** | 15,766 (10.3) | 4,214 (12.6) |
|  | **85+** | 6,676 (4.4) | 2,236 (6.7) |
| **Estimation**  **Comorbidities (% of population)** | **Neoplastic Disease** | 15,290 (10.0) | 2,960 (8.8) |
|  | **Congestive Heart Failure** | 2,076 (1.4) | 578 (1.7) |
|  | **Cerebrovascular Disease** | 3,099 (2.0) | 797 (2.4) |
|  | **Diabetes mellitus** | 4,118 (2.7) | 1,023 (3.1) |
|  | **Pulmonary Disease** | 4,665 (3.1) | 1,520 (4.5) |
| **Migration Background ^2^(% of total)** | **The Netherlands** | 107,881 (70.7) | 24,479 (73.1) |
|  | **Middle & Eastern Europe** | 2,734 (1.8) | 645 (1.9) |
|  | **Other Europe** | 8,224 (5.4) | 1,832 (5.5) |
|  | **Turkey** | 3,470 (2.3) | 729 (2.2) |
|  | **Morocco** | 3,924 (2.6) | 847 (2.5) |
|  | **Suriname** | 7,462 (4.9) | 1,357 (4.1) |
|  | **Dutch Caribbean** | 1,749 (1.2) | 292 (0.9) |
|  | **Indonesia** | 6,227 (4.1) | 1,286 (3.8) |
|  | **Other Africa** | 2,332 (1.5) | 479 (1.4) |
|  | **Other Asia** | 6,596 (4.3) | 1,183 (3.5) |
|  | **Other America’s and Oceania** | 2,005 (1.3) | 358 (1.1) |
| **Socioeconomic status in**  **quintiles ^3^** | **1 (highest)** | 35,810 (23.5) | 7,751 (23.1) |
|  | **2** | 33,200 (21.8) | 6,841 (20.4) |
|  | **3** | 29,876 (19.6) | 6,566 (19.6) |
|  | **4** | 28,689 (18.8) | 6,716 (20.1) |
|  | **5 (lowest)** | 24,239 (15.9) | 5,323 (15.9) |
| **Current smoking (% of population)** | | 36,635 (24.0) | 8,991 (26.9) |
| **Hospitalisation in the past year (% of population)** | | 14,720 (9.7) | 2,959 (8.8) |
| **Clinical diagnosis of pneumonia (% of population)** | | 13,610 (8.9) | 13,172 (39.3) |
| **Same-day corticosteroid prescription with pre-existing asthma or COPD exacerbation (% of population)** | | 45 (0.0) | 105 (0.3) |
| **Current use of systemic corticosteroids (% of population)** | | 1,999 (1.3) | 705 (2.1) |
| **Antibiotics prescribed within 30 days before GP consultation (% of population)** | | 6,110 (4.0) | 1,264 (3.8) |

1. Sex: 1 missing value 2. Migration background: 1 missing variables in the no antibiotics cohort, 2 missing in antibiotics cohort 3. SES: Socioeconomic status on household level, based on financial welfare (standardized income + standardized wealth of household) 791 (0.5%) missing in no antibiotics cohort, 292 (0.9%) missing in antibiotics cohort.

**Table S2: Population Characteristics Validation dataset**

| **Variables** | | **No complications** | **Complications** |
| --- | --- | --- | --- |
| **Subjects n** | | 25,171 | 585 |
| **Sex^1^** | **Female (%)** | 14,855 (59.0) | 283 (48.4) |
|  | **Male (%)** | 10,318 (41.0) | 300 (51.0) |
| **Age in years (% of population)** | **Median (IQR)** | 57  (40 : 71) | 74  (61 : 83) |
|  | **18-49** | 9,904 (39.4) | 91 (15.6) |
|  | **50-64** | 6,397 (25.4) | 85 (14.5) |
|  | **65-74** | 4,404 (17.5) | 122 (20.9) |
|  | **75-84** | 3,306 (13.1) | 182 (31.1) |
|  | **85+** | 1,160 (4.6) | 105 (18.0) |
| **Estimation**  **Comorbidities (% of population)** | **Neoplastic Disease** | 3,888 (15.5) | 131 (22.4) |
|  | **Congestive Heart Failure** | 463 (1.8) | 52 (8.9) |
|  | **Cerebrovascular Disease** | 895 (3.6) | 52 (8.9) |
|  | **Diabetes mellitus** | 1,039 (4.1) | 45 (7.7) |
|  | **Pulmonary Disease** | 1,153 (4.6) | 54 (9.2) |
| **Migration Background ^2^(% of total)** | **The Netherlands** | 17,118 (68.0) | 449 (76.8) |
|  | **Middle & Eastern Europe** | 548 (2.2) | <10 |
|  | **Other Europe** | 1,441 (5.7) | 26 (4.4) |
|  | **Turkey** | 674 (2.7) | 15 (2.6) |
|  | **Morocco** | 752 (3.0) | 13 (2.2) |
|  | **Suriname** | 1,230 (4.9) | 22 (3.8) |
|  | **Dutch Caribbean** | 292 (1.2) | <10 |
|  | **Indonesia** | 930 (3.7) | 21 (3.6) |
|  | **Other Africa** | 462 (1.8) | <10 |
|  | **Other Asia** | 1,342 (5.3) | 15 (2.6) |
|  | **Other America’s and Oceania** | 382 (1.5) | <10 |
| **Socioeconomic status in**  **quintiles ^3^** | **1 (highest)** | 5,670 (22.5) | 89 (15.2) |
|  | **2** | 5,363 (21.3) | 106 (18.1) |
|  | **3** | 5,008 (19.9) | 117 (20.0) |
|  | **4** | 4,683 (18.6) | 138 (23.6) |
|  | **5 (lowest)** | 4,272 (17.0) | 133 (22.7) |
| **Current smoking (% of population)** | | 5,910 (23.5) | 169 (28.9) |
| **Hospitalisation in the past year (% of population)** | | 2,348 (9.3) | 180 (30.8) |
| **Working diagnosis of pneumonia (% of population)** | | 3,699 (14.7) | 245 (41.9) |
| **Same-day corticosteroid prescription with pre-existing asthma or COPD exacerbation (% of population)** | | 35 (0.1) | <10 |
| **Current use of systemic corticosteroids (% of population)** | | 456 (1.8) | 47 (8.0) |
| **Antibiotics prescribed within 30 days before GP consultation (% of population)** | | 1,227 (4.9) | 93 (15.9) |
| **Same-day antibiotics prescription**  **(% of population)** | | 4,715 (18.7) | 173 (29.6) |

1. SES: Socioeconomic status on household level, based on financial welfare (standardized income + standardized wealth of household) 175 (0.7%) missing in no complications cohort, 2 (0.3%) missing complications cohort.

**Table S3: Difference in deviance and deviance ratio between models**

|  | **Sample** | **Deviance** | **Deviance ratio** |
| --- | --- | --- | --- |
| **Model 1 without SES and Migration background** | Training | 0.184 | 0.1216 |
|  | Testing | 0.191 | 0.1225 |
| **Model 2 with SES and Migration background** | Training | 0.184 | 0.1233 |
|  | Testing | 0.190 | 0.1242 |
| **Model 3 with SES and Migration background** | Training | 0.184 | 0.1236 |
|  | Testing | 0.190 | 0.1244 |

**Table S4: Absolute risk and predicted risk of a patient’s risk of developing a complicated LRTI classified into risk-groups according to NHG guidelines, stratified by SES-quintiles in derivation dataset**

|  | | **Absolute risk** | **Predicted risk** |
| --- | --- | --- | --- |
| **Risk strata 1** | **1^st^ quintile** | 0.7 | 0.9 |
|  | **2^nd^ quintile** | 0.8 | 0.9 |
|  | **3^rd^ quintile** | 0.8 | 1.0 |
|  | **4^th^ quintile** | 0.8 | 1.1 |
|  | **5^th^ quintile** | 1.0 | 1.1 |
| **Risk strata 2** | **1^st^ quintile** | 1.9 | 1.7 |
|  | **2^nd^ quintile** | 1.9 | 1.7 |
|  | **3^rd^ quintile** | 2.2 | 2.1 |
|  | **4^th^ quintile** | 3.2 | 2.8 |
|  | **5^th^ quintile** | 2.6 | 2.5 |
| **Risk strata 3** | **1^st^ quintile** | 5.0 | 4.9 |
|  | **2^nd^ quintile** | 5.1 | 5.0 |
|  | **3^rd^ quintile** | 6.4 | 6.2 |
|  | **4^th^ quintile** | 8.7 | 8.9 |
|  | **5^th^ quintile** | 7.2 | 7.5 |

**Table S5: Absolute risk and predicted risk of a patient’s risk of developing a complicated LRTI classified into risk-groups according to NHG guidelines stratified by SES-quintiles in validation dataset**

|  | | **Absolute risk** | **Predicted risk** |
| --- | --- | --- | --- |
| **Risk strata 1** | **1^st^ quintile** | 0.4 | 0.9 |
|  | **2^nd^ quintile** | 0.5 | 0.9 |
|  | **3^rd^ quintile** | 0.7 | 1.0 |
|  | **4^th^ quintile** | 0.6 | 1.1 |
|  | **5^th^ quintile** | 1.1 | 1.1 |
| **Risk strata 2** | **1^st^ quintile** | 2.0 | 1.9 |
|  | **2^nd^ quintile** | 2.7 | 2.0 |
|  | **3^rd^ quintile** | 2.3 | 2.3 |
|  | **4^th^ quintile** | 3.3 | 3.1 |
|  | **5^th^ quintile** | 3.3 | 3.1 |
| **Risk strata 3** | **1^st^ quintile** | 4.3 | 4.8 |
|  | **2^nd^ quintile** | 4.9 | 5.2 |
|  | **3^rd^ quintile** | 7.4 | 6.2 |
|  | **4^th^ quintile** | 7.8 | 8.2 |
|  | **5^th^ quintile** | 7.3 | 8.9 |

**Table S6:** Increase in mean predicted probability of a complicated course of LRTI across NHG risk strata, adjusted for all model covariates

| NHG | | **Marginal effect %** | **Upper boundary % 95%CI** | **Lower boundary %**  **95%CI** |
| --- | --- | --- | --- | --- |
| **Risk strata 1** | **1^st^ quintile** | REF | REF | REF |
|  | **2^nd^ quintile** | 0.09 | 0.09 | 0.09 |
|  | **3^rd^ quintile** | 0.21 | 0.21 | 0.21 |
|  | **4^th^ quintile** | 0.28 | 0.28 | 0.28 |
|  | **5^th^ quintile** | 0.36 | 0.36 | 0.36 |
| **Risk strata 2** | **1^st^ quintile** | REF | REF | REF |
|  | **2^nd^ quintile** | 0.19 | 0.19 | 0.18 |
|  | **3^rd^ quintile** | 0.45 | 0.46 | 0.45 |
|  | **4^th^ quintile** | 0.60 | 0.60 | 0.60 |
|  | **5^th^ quintile** | 0.76 | 0.77 | 0.76 |
| **Risk strata 3** | **1^st^ quintile** | REF | REF | REF |
|  | **2^nd^ quintile** | 0.52 | 0.52 | 0.51 |
|  | **3^rd^ quintile** | 1.26 | 1.27 | 1.25 |
|  | **4^th^ quintile** | 1.66 | 1.68 | 1.64 |
|  | **5^th^ quintile** | 2.10 | 2.11 | 2.08 |

**Table S7: Multivariable logistic regression sub analyses with CRP added in the model, only practices where a CRP was possible are included, prediction models refined through LASSO selection**

| **Independent Variables** | | **Refined full multivariable conventional model (1)**  **OR (95% CI)** | **Refined full multivariable SES model (2)**  **OR (95% CI)** | **Refined full multivariable migration model (3)**  **OR (95% CI)** |
| --- | --- | --- | --- | --- |
| **Sex** | |  |  |  |
| Male | | **Ref** | **Ref** | **Ref** |
| Female | | 0.89 (0.82 – 0.96) | 0.87 (0.81 – 0.94) | 0.87 (0.81 – 0.94) |
| **Age** | |  |  |  |
| 18 – 49 | | **Ref** | **Ref** | **Ref** |
| 50 – 64 | | 1.40 (1.23 – 1.59) | 1.46 (1.28 – 1.67) | 1.44 (1.27 – 1.64) |
| 65 – 74 | | 2.50 (2.21 – 2.84) | 2.59 (2.29 – 2.94) | 2.57 (2.26 – 2.91) |
| 75 – 84 | | 3.52 (3.09 – 4.00) | 3.53 (3.10 – 4.01) | 3.51 (3.08 – 4.00) |
| 85+ | | 6.60 (5.75 – 7.58) | 6.54 (5.69 – 7.51) | 6.50 (5.64 – 7.49) |
| **SES (quintiles where 5 is the lowest)** | |  |  |  |
| 1 | | - | **Ref** | **Ref** |
| 2 | | - | 1.12 (0.99 – 1.27) | 1.12 (0.99 – 1.27) |
| 3 | | - | 1.26 (1.12 – 1.43) | 1.26 (1.11 – 1.42) |
| 4 | | - | 1.34 (1.19 – 1.50) | 1.33 (1.18 – 1.50) |
| 5 | | - | 1.46 (1.29 – 1.66) | 1.44 (1.27 – 1.64) |
| **Migration Background** | |  |  |  |
| The Netherlands | | - | - | **Ref** |
| Middle & Eastern Europe | | - | - | 0.48 (0.28 – 0.82) |
| Other Europe | | - | - | 0.90 (0.75 – 1.07) |
| Turkey | | - | - | 1.16 (0.88 – 1.53) |
| Morocco | | - | - | 1.11 (0.87 – 1.43) |
| Suriname | | - | - | 1.01 (0.83 – 1.23) |
| Dutch Caribbean | | - | - | 1.16 (0.80 – 1.70) |
| Indonesia | | - | - | 1.05 (0.88 – 1.26) |
| Other Africa | | - | - | 1.12 (0.78 – 1.60) |
| Other Asia | | - | - | 0.96 (0.75 – 1.21) |
| Other America’s and Oceania | | - | - | 0.92 (0.60 – 1.43) |
| **Comorbidities** | |  |  |  |
| Neoplastic Disease | | 1.60 (1.45 – 1.77) | 1.62 (1.47 – 1.79) | 1.62 (1.47 – 1.79) |
| Congestive Heart Failure | | 1.80 (1.54 – 2.11) | 1.78 (1.52 – 2.09) | 1.78 (1.52 – 2.09) |
| Cerebrovascular Disease | | 1.17 (1.00 – 1.38) | 1.17 (0.99 – 1.38) | 1.17 (0.99 – 1.38) |
| Diabetes mellitus | | 1.89 (1.09 – 3.29) | 1.82 (1.05 – 3.17) | 1.79 (1.03 – 3.11) |
| Pulmonary disease | | 1.18 (1.01 – 1.39) | 1.16 (0.99 – 1.36) | 1.16 (0.99 – 1.37) |
| **Health** | |  |  |  |
| Current smoking | | 1.10 (1.00 – 1.20) | 1.06 (0.97 – 1.16) | 1.07 (0.98 – 1.17) |
| Hospitalisation in the past year | | 2.07 (1.88 – 2.28) | 2.03 (1.84 – 2.23) | 2.03 (1.84 – 2.23) |
| Current use of oral systemic corticosteroids | | 2.10 (1.77 – 2.49) | 2.09 (1.76 – 2.48) | 2.08 (1.76 – 2.47) |
| Antibiotics prescribed <30 days before GP consultation | | 1.38 (1.20 – 1.57) | 1.37 (1.20 – 1.57) | 1.37 (1.20 – 1.57) |
| **Degree of illness** | |  |  |  |
| CRP-POC-test | Unknown/not done | 1.11 (0.57 – 2.15) | 1.13 (0.58 – 2.19) | 1.14 (0.59 – 2.21) |
|  | 0-20 | **Ref** | **Ref** | **Ref** |
|  | 20-100 | 0.92 (0.28 – 3.02) | 0.94 (0.29 – 3.10) | 0.94 (0.29 – 3.``) |
|  | >100 or diagnosis of pneumonia | 3.18 (1.63 – 6.19) | 3.24 (1.66 – 6.32) | 3.28 (1.68 – 6.38) |
| Same-day antibiotics prescription | | 1.00 (0.91 – 1.10) | 1.00 (0.90 – 1.10) | 1.00 (0.91 – 1.10) |
| Same-day corticosteroid prescription with pre-existing asthma or COPD | | 1.48 (0.74 – 2.97) | 1.50 (0.75 – 3.01) | 1.50 (0.75 – 3.01) |
| **Interactions** | |  | | |
| Age category 18-49* Diabetes Mellitus | | **Ref** | **Ref** | **Ref** |
| Age category 50-64* Diabetes Mellitus | | 0.87 (0.46 – 1.64) | 0.86 (0.46 – 1.62) | 0.87 (0.46 – 1.64) |
| Age category 65-74* Diabetes Mellitus | | 0.67 (0.35 – 1.27) | 0.66 (0.35 – 1.25) | 0.67 (0.35 – 1.27) |
| Age category 75-84* Diabetes Mellitus | | 0.60 (0.31 – 1.15) | 0.60 (0.31 – 1.15) | 0.61 (0.31 – 1.17) |
| Age category >85* Diabetes Mellitus | | 0.33 (0.15 – 0.71) | 0.34 (0.16 – 0.73) | 0.35 (0.16 – 0.74) |

All models were refined through LASSO selection procedures. Population includes only individuals at a GP-practice where a CRP point of care test could be conducted.

**Table S8: difference in discrimination between models with CRP-POC test integrated**

| Derivation dataset | **Model 1 without SES and Migration background** | **Model 2 with SES** | **Model 3 with SES and Migration background** |
| --- | --- | --- | --- |
| **AUROC (95% CI)** | 0.784 (0.774 – 0.793) | 0.787 (0.778 – 0.796) | 0.787 (0.778 – 0.797) |
| **ROC-curve bootstrap** | 0.778 (0.769 – 0.787) | 0.785 (0.777 – 0.795) | 0.786 (0.778 – 0.795) |
| **Brier-score** | 0.020 | 0.020 | 0.020 |
| **AIC** | 25045.11 | 25001.22 | 24991.92 |
| Validation dataset | **Model 1 without SES and Migration background** | **Model 2 with SES** | **Model 3 with SES and Migration background** |
| **AUROC (95% CI)** | 0.781 (0.764 – 0.806) | 0.786 (0.770 – 0.810) | 0.786 (0.770 – 0.811) |
| **ROC-curve bootstrap** | 0.777 (0.754 – 0.798) | 0.786 (0.763 – 0.806) | 0.786 (0.764 – 0.807) |
| **Brier-score** | 0.021 | 0.021 | 0.021 |
| **AIC** | 4886.68 | 4876.492 | 4875.233 |

**Table S9: Difference in Calibration between models with CRP-POC test integrated**

| Derivation dataset | **Model 1 without SES and Migration background** | **Model 2 with SES** | **Model 3 with SES and Migration background** |
| --- | --- | --- | --- |
| **Intercept (95% CI)** | 0.07 (-0.05 – 0.18) | 0.07 (-0.05 – 0.18) | 0.07 (-0.04 – 0.19) |
| **Slope (95% CI)** | 1.020 (0.988 – 1.052) | 1.020 (0.988 – 1.052) | 1.022 (0.990 – 1.054) |
| **Brier-score** | 0.020 | 0.020 | 0.020 |
| Validation dataset | **Model 1 without SES and Migration background** | **Model 2 with SES** | **Model 3 with SES and Migration background** |
| **Intercept (95% CI)** | 0.02 (-0.24 – 0.27) | 0.00 (-0.24 – 0.25) | 0.01 (-0.24 – 0.26) |
| **Slope (95% CI)** | 1.014 (0.940 – 1.087) | 1.012 (0.939 – 1.085) | 1.014 (0.941 – 1.086) |
| **Brier-score** | 0.021 | 0.021 | 0.021 |

**Table S10: Difference in deviance and deviance ratio between models with CRP-POC test integrated**

|  | **Sample** | **Deviance** | **Deviance ratio** |
| --- | --- | --- | --- |
| **Model 1 without SES and Migration background** | Training | 0.181 | 0.1235 |
|  | Testing | 0.191 | 0.1220 |
| **Model 2 with SES and Migration background** | Training | 0.181 | 0.1250 |
|  | Testing | 0.191 | 0.1238 |
| **Model 3 with SES and Migration background** | Training | 0.183 | 0.1253 |
|  | Testing | 0.190 | 0.1240 |

**Figure F1: Calibration plot conventional model**

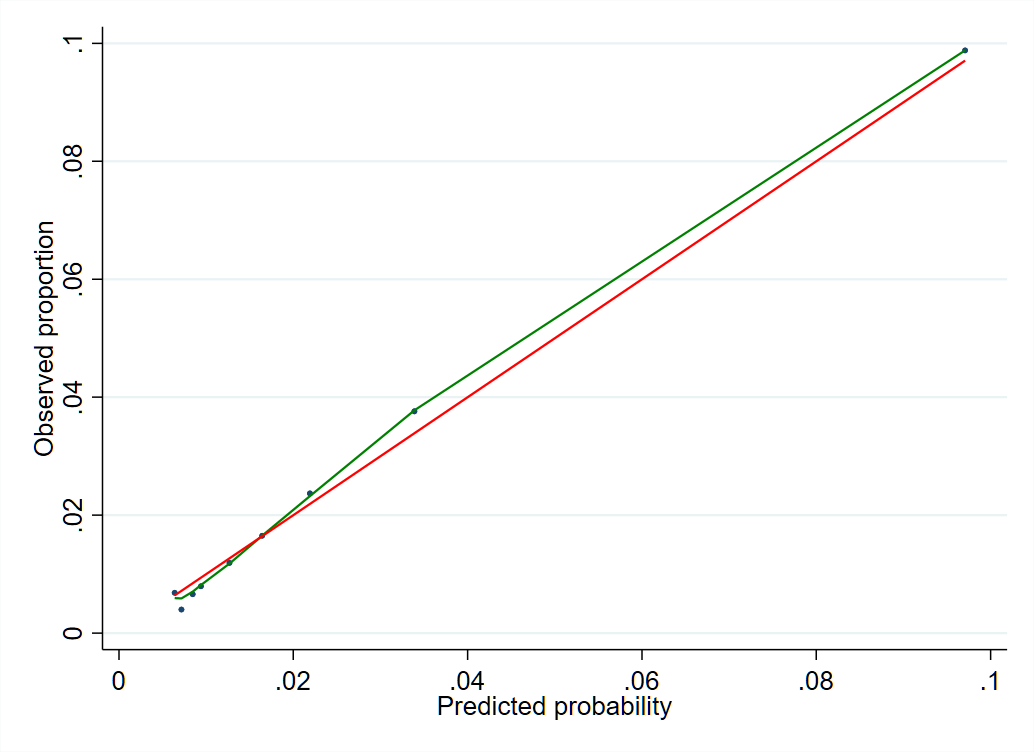

**Figure F2: Calibration plot SES model**

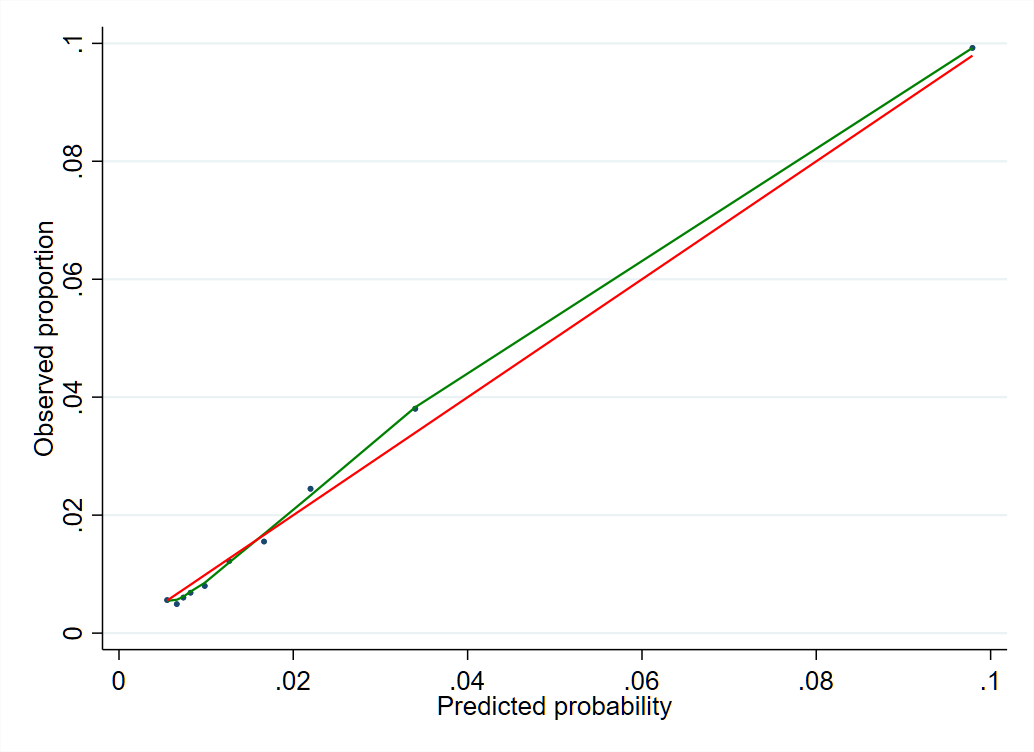

**Figure F3: Calibration plot migration model**

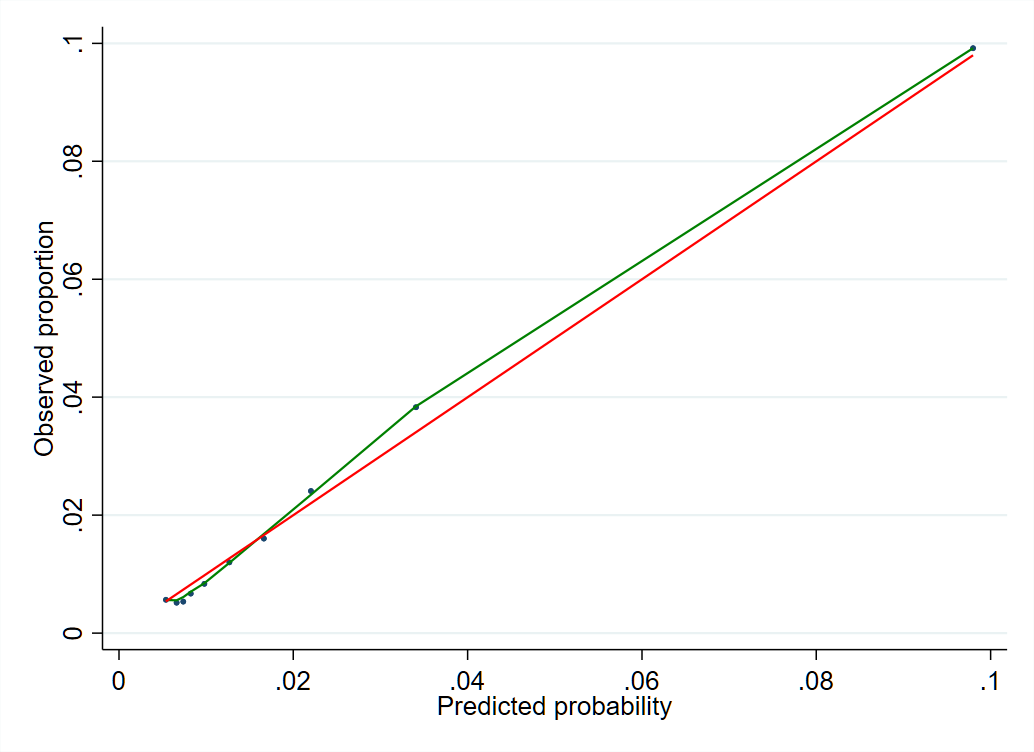

**M-references**

1. Fine MJ, Auble TE, Yealy DM, et al. A prediction rule to identify low-risk patients with community-acquired pneumonia. N Engl J Med. 1997;336(4):243-50.
2. World Health Organization. Anatomical Therapeutic Chemical (ATC) Classification. <https://www.who.int/tools/atc-ddd-toolkit/atc-classification>. Date last accessed: December 4 2024
